# A Comparison of Methylprednisolone and Dexamethasone in Intensive Care Patients with COVID-19

**DOI:** 10.1101/2021.02.03.21251088

**Authors:** Justine J. Ko, Clay Wu, Neha Mehta, Noah Wald-Dickler, Wei Yang, Renli Qiao

## Abstract

**OBJECTIVES:** This study retrospectively compares the effectiveness of methylprednisolone to dexamethasone in patients with severe acute respiratory syndrome coronavirus 2 (SARS-CoV-2 or COVID-19) requiring ICU care.

**DESIGN:** This is an institutional review board approved cohort study in patients with COVID-19 requiring intensive care unit admission. Patients admitted and requiring oxygen supplementation were treated with either methylprednisolone or dexamethasone.

**SETTING:** This study takes place in the intensive care units at a large, tertiary, public teaching hospital serving a primarily low-income community in urban Los Angeles.

**PATIENTS:** All eligible patients admitted to the intensive care unit for COVID-19 respiratory failure from March 1 to July 31, 2020 were included in this study.

**INTERVENTIONS:** A total of 262 patients were grouped as receiving usual care (n=75), methylprednisolone dosed at least at 1mg/kg/day for ≥ 3 days (n=104), or dexamethasone dosed at least at 6 mg for ≥ 7 days (n=83).

**MEASUREMENTS and MAIN RESULTS:** All-cause mortality within 50 days of initial corticosteroid treatment as compared to usual care was calculated. The mortality effect was then stratified based on levels of respiratory support received by the patient.

In this cohort of 262 patients with severe COVID-19, all-cause mortalities in the usual care, methylprednisolone, and dexamethasone groups were 41.3%, 16.4% and 26.5% at 50 days (p <0.01) respectively. In patients requiring mechanical ventilation, mortality was 42% lower in the methylprednisolone group than in the dexamethasone group (hazard ratio 0.48, 95% CI: 0.235-0.956, p=0.0385).

**CONCLUSIONS:** In COVID-19 patients requiring mechanical ventilation, sufficiently dosed methylprednisolone can lead to a further decreased mortality as compared to dexamethasone.

## INTRODUCTION

The coronavirus disease 2019 (COVID-19) continues to be an aggressive pandemic.^1-3^ Approximately 5% patients that develop respiratory failure require intensive care unit (ICU) level of care and are associated with a high mortality.^4^ The pathophysiology of COVID-19 is primarily attributed to excessive inflammatory reaction.^5^ Recently, the RECOVERY trial proved a mortality benefit for use of dexamethasone in severe disease.^6^ Publication of this trial has led WHO to recommend systemic corticosteroids in patients with severe COVID-19 and halted other trials.^7^

However, it has been well established that among various corticosteroids, methylprednisolone, but not dexamethasone, is the clinically preferred agent for its anti-inflammatory effect and high dose is required to achieve the therapeutic effect of the steroid.^8-11^ As such, questions remain: Is the effect seen with dexamethasone drug-specific, or is it an anti-inflammatory effect of corticosteroid in general? If so, can sufficiently dosed methylprednisolone provide further benefit? Unfortunately, in the meta-analysis used for corticosteroid recommendations, data on methylprednisolone was limited and it is unlikely prospective trials can be conducted timely for such a comparison.^7^

Here, we report our observation since the beginning of the pandemic. We have compared the effectiveness of methylprednisolone vs dexamethasone in a large cohort.

## MATERIALS AND METHODS

### Experiment Design

In March 2020, we originally proposed a course of high-dose, short-term methylprednisolone to prospectively treat COVID-19 patients with severe hypoxemia requiring admission to ICU. However, it was up to the treating physicians to decide individual plan and we were able to convince only some colleagues to use steroid. Patients who were not treated with steroid constituted the usual care group. Due to abrupt surge of COVID-19 cases leading to overwhelming workload and delay in IRB approval, the study design was changed to retrospective analysis. The proposal was submitted in May 15, 2020 and was approved by the institutional review board (IRB) of University of Southern California (USC) on July 20, 2020.

Following publication of the RECOVERY trial, dexamethasone became a standard care. No more patients was added to the usual care group since, but our team continued to rely on the methylprednisolone regimen as a stronger benefit from methylprednisolone compared to dexamethasone has become clear in our preliminary results by this time. The study proposal was then modified to compare the mortalities among 3 groups, usual care (no steroid treatment), methylprednisolone, and dexamethasone. All the modifications in the study protocol were approved by IRB of USC.

Data was collected between March 1 to July 31, 2020 at a large, public hospital. The observation days were counted from the day a steroid was started or the day of ICU admission in the usual care group. All patients were tested for COVID-19 upon admission via a nucleic acid assay.

### Study Population

All patients who tested positive and were admitted to ICU for respiratory failure from COVID-19 during the collection period were evaluated. Criteria for ICU admission for COVID-19 were an oxygen requirement of ≥ 40 L/min and FiO_2_ of ≥ 50%.

Exclusion criteria included patients in whom COVID-19 was an incidental finding.. The most common reasons for these ICU admissions were: trauma and other surgical conditions, diabetic ketoacidosis (DKA) requiring insulin drips, drug/alcohol intoxication, cardiac events, non-COVID-19 related sepsis, and GI bleeds requiring endoscopy. Of the 285 ICU admissions, the total number of cases in the final analysis was 262 (92% of total).

### Treatment Groups

The study compared the outcome of three groups of patients with minimum dosing as following: 1) methylprednisolone 1 mg/kg/day for ≥ 3 days, 2) dexamethasone 6 mg/day for ≥ 7 days, and 3) usual care only. The usual care per our hospital’s guidelines included the following: 1) anticoagulation based on D-Dimer levels (see below); 2) prone positioning when PF ratio is <150 (16 hours prone alternating with 8 hours supine for patients on mechanical ventilation; whenever feasible in patients on non-invasive oxygen supplement); 3) antibiotics for 5 days if procalcitonin ≥ 0.25 ng/mL on admission. Some COVID-19 patients were started on remdesivir or convalescent plasma and these effects were analyzed. Very few patients received hydroxychloroquine or tocilizulamab and these numbers were insufficient for statistical analysis. No patient required ECMO support.

The anticoagulation protocol used in our hospital is as following. For D-Dimer <6 FEU/ml, start heparin product at prophylactic dosing for DVT. For D-Dimer >6 or increase by 2, low therapeutic dosing. If evidence of VTE is present, start full anticoagulation using heparin products. For VTE despite therapeutic dosing of heparin, use anti-Xa agents or argatroban or fondaparinux. Full version of our anticoagulation protocol is attached.

### Outcome Measures

We report an all-cause mortality within 50 days of initial treatment as the primary outcome. Secondary outcomes included ICU length of stay and 28^th^ day mortality.

### Statistical Analysis

The hazard ratio from Cox Regression was conducted to estimate the case fatality rate ratio (RR) and the cut off was set at 50 days from symptom onset. Kaplan–Meier survival curves were constructed to show cumulative mortality over the 50-day period. Chi-square tests were used for the different mortality rates among subject characteristics and other related risk factors. The RRs were stratified based on requirement of mechanical ventilation when comparing outcomes between treatments. All p-values were two-sided and p <0.05 was considered significant.

## RESULTS

### Patient Characteristics

The baseline demographic data is summarized in Table 1. Most patients were of Hispanic race (82%). The overall mean (±SD) age of these patients was 57.8±14.5 years with 44% above 60 years. 76% of the patients were male. These patients had a mean BMI of 30.6 ±7.9. In terms of their comorbidities, 22.8% had hypertension, 37.2% had diabetes, 7.3% had heart disease, 6.7% had chronic lung disease, 9.1% had chronic kidney disease, 3.5% had liver disease, and 4.9% had underlying malignancy, with 55% of the patients carrying at least one of these diagnoses. The demographic characteristics were comparable among the three groups (Table 1a). The average days on steroid were 8.12±3.29 days of which 78% was 10 days in the dexamethasone group. The average days on steroid were 5.25±3.57 days of which 75% was 3-6 days in the methylprednisolone group.

**Table 1a.** Demographic data by treatment groups

|  | Usual care<br>(n=75) | Dexamethasone<br>(n=83) | Methylprednisone<br>(n=104) |
| --- | --- | --- | --- |
| Age |  |  |  |
| Mean±SD (years) | 60.5±15.3 | 57.8±14.9 | 56.2±14.1 |
| % <60 yo | 63% | 53% | 58% |
| % ≥60 yo | 37% | 47% | 42% |
| Sex ( %) |  |  |  |
| Male | 75% | 75% | 76% |
| Female | 25% | 25% | 24% |
| Race (%) |  |  |  |
| White | 3% | 0 | 3% |
| AA | 4% | 2% | 2% |
| Hispanic | 77% | 89% | 81% |
| Asians | 11% | 4% | 4% |
| Others | 5% | 6% | 11% |
| BMI (kg/m<sup>2</sup>)<br>(mean±SD) | 29.5±8.8 | 30.8± 7.9 | 31.2± 7.5 |
| Comorbidities (%) |  |  |  |
| Hypertension | 13.3% | 28.2% | 19.0% |
| DM | 43.4% | 34.1% | 34.8% |
| Heart diseases | 6.6% | 8.2% | 6.7% |
| Kidney Diseases | 9.3% | 7.1% | 10.3% |
| Lung diseases | 6.6% | 11.8% | 9.2% |
| HIV | 0 | 1.2% | 0 |
| Malignancy | 3.9% | 8.2% | 2.9% |
| Liver diseases | 5.2% | 7.4% | 9.2% |
| Other Treatment<br>Received (n, %) |  |  |  |
| Remdesivir | 8 (11%) | 21 (25%) | 23 (22%) |
| Convalescent Plasma | 19 (25%) | 22 (27%) | 31 (30%) |
| Hydroxychloroquine | 6 | 2 | 3 |
| Tocilizumab | 0 | 0 | 2 |
| Days on steroid |  | 8.12±3.29 | 5.25±3.57 |

**Table 1b.** Severity of hypoxemia

|  | Usual Care | Dexamethasone | Methylprednisolone |
| --- | --- | --- | --- |
| Mechanical Ventilation (%) | 57.8% | 43.5% | 45.7% |
| Lowest PF Ratio (Mean±SD) | 99.1±45.5 | 107.8±74 | 88.17±47.1 |
| Highest PEEP (cmH<sub>2</sub>O) (Mean±SD) | 10.2±6.1 | 9.2±5.6 | 9.5±6.7 |

No significant difference among the 3 groups in all the parameters. PF Ratio = PaO<sub>2</sub>/FiO<sub>2</sub>. For analysis of the highest PEEP, PEEP was assumed to be 5 cmH<sub>2</sub>O in patients on HFNC oxygen supplement.

The severity of hypoxemia was described by requirement of mechanical ventilation, worst PaO_2_/FiO_2_ ratio (PF Ratio), and highest PEEP (Positive End Expiration Pressure, cmH_2_O) required. In patients on high flow nasal cannula (HFNC) for oxygen supplement, 5 cmH_2_O was used as PEEP. Mechanical ventilation was required to maintain oxygenation in 55% patients, while the rest were on HFNC. The lowest PF ratios were 99.1±45.5, 107.8±74 and 88.17±47.1 (P>0.05) and the highest PEEP required were 10.2±6.1, 9.2±5.6, 9.5±6.7 cmH_2_O (P>0.05) (Table 1b) in the usual care, dexamethasone and methylprednisolone groups respectively (Table 1b), indicating that all patients from the 3 different treatment groups suffered from similar severity of hypoxemia due to COVID-19.

In Table 2, all data from the 3 groups were pooled to analyze the potential impact of each parameter on mortality. Older age, preexistence of any co-morbidity, and requirement of mechanical ventilation were identified as risk factors for increased mortality (Table 2). However, these potential confounding factors were distributed in similar pattern among the 3 groups (Table 1).

**Table 2.** Case Fatality Rate by Characteristics and Treatments in All the Patients From the 3 groups

| Factors |  | % | P |
| --- | --- | --- | --- |
| Total (N=262) |  | 26.72 |  |
| Age | <60 | 16.44 | <.0001 <sup>a</sup> |
|  | ≥ 60 | 39.66 |  |
| Sex | Male | 23.74 | 0.0552 |
|  | Female | 35.94 |  |
| Race/Ethnicity | White | 20 | 0.9446 |
|  | Black | 28.57 |  |
|  | Hispanics | 26.17 |  |
|  | Asians | 36.36 |  |
|  | Others | 29.17 |  |
| BMI | <30 | 31.08 | 0.069 |
|  | ≥30 | 21.05 |  |
| Pre-existing Medical Condition | No | 18.8 | 0.0093 <sup>a</sup> |
|  | Yes | 33.1 |  |
| Mechanic Ventilation | No | 11.21 | <.0001 <sup>a</sup> |
|  | Yes | 42.97 |  |
| Remdesivir | No | 26.7 | 0.9923 |
|  | Yes | 26.76 |  |
| Convalescent Plasma | No | 26.04 | 0.6822 |
|  | Yes | 28.57 |  |

<sup>a</sup> older age (<60 vs ≥ 60 years), pre-existence of any medical conditions, and requirement of mechanical ventilation showed significant impacts on case fatality rate.

Some patients received other agents proposed to be potentially effective for COVID-19. RR analysis showed no statistically significant effect by either remdesivir or convalescent plasma on outcome (Table 2).

### Primary Outcome

The overall mortality in the entire cohort was 26.7%. Mortality at 50^th^ day was 41.3% in usual care, 26.5% in dexamethasone, and 16.4% in methylprednisolone, statistically significantly lower in both steroid groups compared to usual care (Table 3, p<0.001). The mortality and the magnitude of its improvement with dexamethasone compared to usual care matched that reported in the RECOVERY trial (26.5% vs 41.3%, a 34% reduction) (6). After adjusting for 7 confounding factors, no significant difference in mortality benefit was identified between the methylprednisolone and the dexamethasone groups (Figure 1) (RR 0.635, 95% CI: 0.326-1.218, p=0.173).

**Table 3.** Adjusted Outcomes by Treatment

| Treatment | Usual Care | Dexamethasone | Methylprednisolone |
| --- | --- | --- | --- |
| Total (n) | 75 | 83 | 104 |
| Primary |  |  |  |
| Deaths at 50<sup>th</sup> day (n) | 31 | 22 | 17 |
| Case fatality at 50<sup>th</sup> Day | 41.3% | 26.5%<sup>A</sup> | 16.4%<sup>A</sup> |
| Secondary |  |  |  |
| Case fatality at 28<sup>th</sup> Day | 35.5% | 23.5%<sup>A</sup> | 15.2%<sup>A</sup> |
| ICU Length of Stay |  |  |  |
| (days, mean ± SD) | 12.6 ± 13.5 | 14.3 ± 13.1 | 13.4 ± 14.8 |

**Figure 1.**
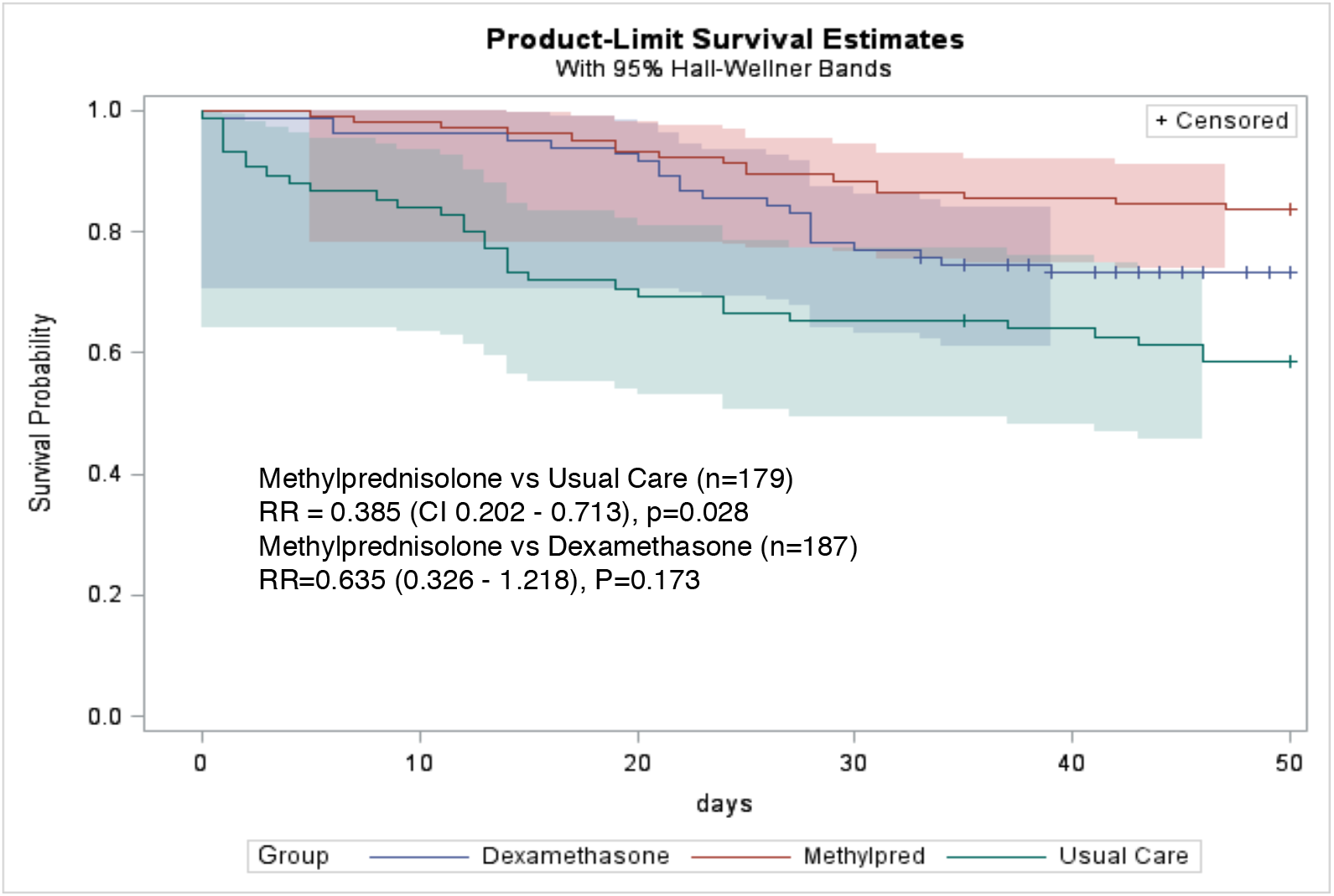
Case Fatality rates at 50th Day. All patients in different intervention groups (n=262) Kaplan–Meier survival curves for 50th-day mortality among all the patients in the study (primary outcome). Both dexamethasone and methylprednisolone confer significant reduction in mortality (P<0.05 for both). The difference between methylprednisolone and dexamethasone groups did not reach statistic significance (P=0.173).

After stratification by level of respiratory support, the mortality was 50.0 % in patients requiring mechanical ventilation and 31.6% in patients on HFNC in the usual care group. In comparison, the mortality in patients requiring mechanical ventilation was 31% in the methylprednisolone group and 54% in the dexamethasone group (Figure 2) (RR 0.480, 95% CI: 0.235-0.956, p=0.0385); however, mortality in patients who did not require mechanical ventilation was 5.3% in the methylprednisolone group and 6.5% in the dexamethasone groups (Figure 3) (RR 0.697, 95% CI: 0.129-3.776, p=0.659). Thus, in patients requiring mechanical ventilation, methylprednisolone was superior to dexamethasone in reducing mortality of COVID-19. However, in patients who did not require mechanical ventilation, both steroids decreased mortality to low rates and no difference between the two steroids could be detected.

**Figure 2.**
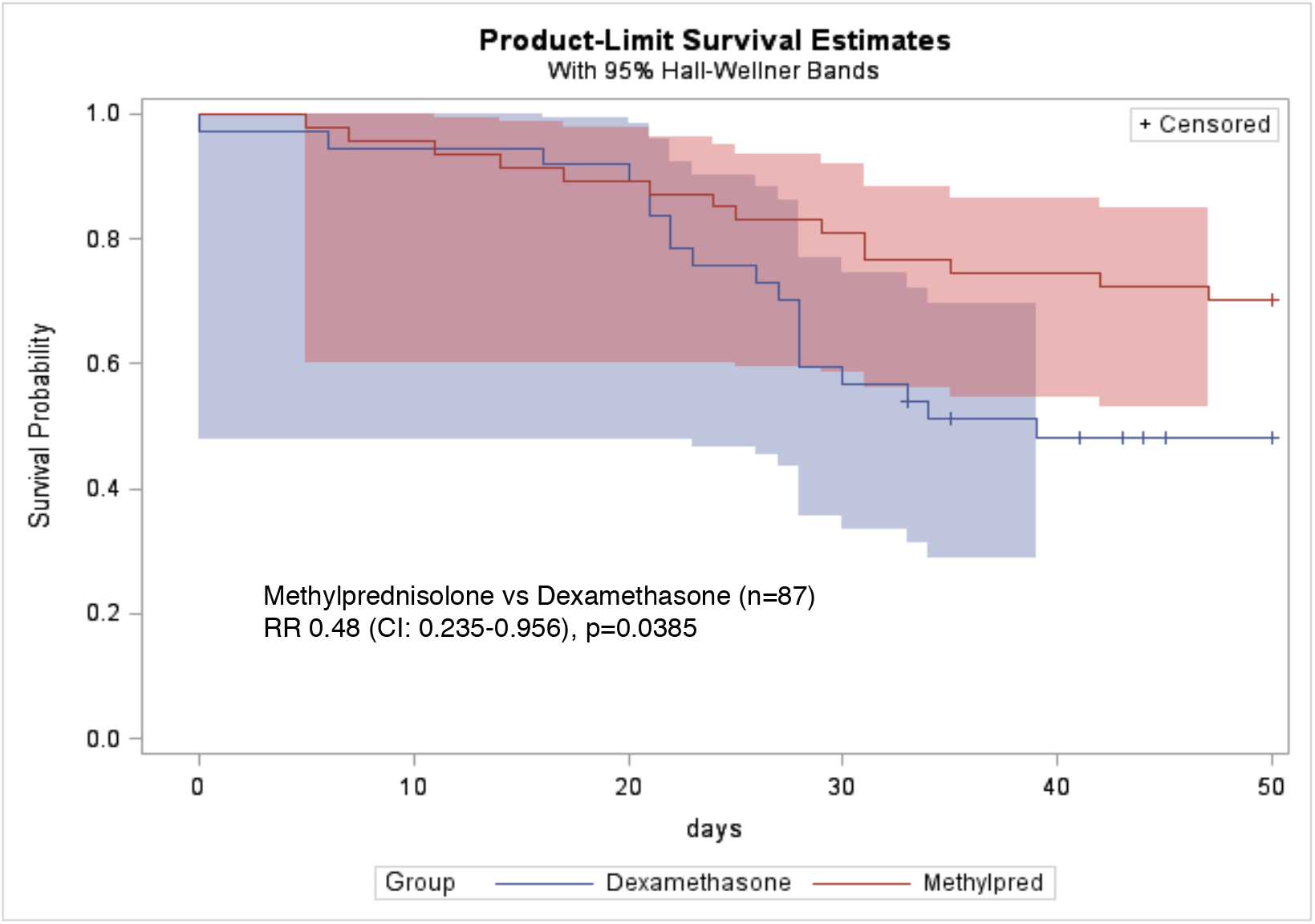
Fatality rate ratio of methylprednisolone (48) vs dexamethasone (39) in patients on mechanical ventilation. Kaplan–Meier survival curves for 50th-day mortality in patients on mechanical ventilation at the time when corticosteroid was started. Further mortality benefit was seen in favor of methylprednisolone over dexamethasone in patients requiring mechanical ventilation (P<0.05).

### Secondary Outcome

The ICU lengths of stay mortality were similar among all three groups (Table 3). The differences in 28^th^ day mortality among the 3 groups were similar to that of 50^th^ day mortality (Table 3).

## DISCUSSION

Corticosteroids are synthetic analogues of the natural steroid that can halt the production of cytokines and thereby confer anti-inflammatory effect^8^. It has been shown that patients with COVID-19 release elevated level of cytokines and autopsy has shown an alveolar damage that is steroid responsive.^12^ The RECOVERY trial has established that dexamethasone can significantly decrease mortality in severe cases of COVID-19, especially in the critical condition when ventilator support is required, presumably because the more severe lung injury was a result of worse hyperinflammation.^6-7^

In the RECOVERY trial, the rationale of using dexamethasone included its anti-inflammatory potency, lack of mineralocorticoid effect, and longer acting profile.^6-7^ However, none of these confers unique advantage for dexamethasone over other steroids, especially in the ICU setting where drug administration is not a barrier. Studies have shown difference of various steroids in their anti-inflammatory biology, beside potency and duration.^8^ For this reason, it is methylprednisolone, but not dexamethasone, that has long been the clinically preferred steroid for an anti-inflammatory effect in lung diseases, such as in acute lung transplant rejection, pneumonitis in various autoimmune conditions, and drug toxicities.^8-11^ A high dose of steroids is usually required in these conditions to achieve a direct effect on cell membrane-associated proteins.^13^ Shorter duration of therapy can minimize side effects.^8^

Based on these knowledge/experience accumulated from treating the above discussed conditions in which the inflammatory and autoimmune injury were also mediated by cytokine release, we implemented a regimen of methylprednisolone at high dose over a short time course from the beginning of the pandemic, prior to publication of the RECOVERY trial. Based on steroid bio-equivalency, the steroid anti-inflammatory potency of our regimen was over 2-fold higher administered in approximately half the duration as compared to use of dexamethasone in the RECOVERY trial.^6,14^

Our results provided evidence that the mortality benefit of dexamethasone in severe COVID-19 is not drug specific but rather, is likely through a general anti-inflammatory effect of corticosteroid. More importantly, the higher anti-inflammatory potency of our methylprednisolone regimen can confer further mortality benefit beyond dexamethasone, especially in patients requiring mechanical ventilation.

A major longitudinal change in the management of COVID-19 cases is the expanded use of HFNC for oxygen supplement. During the very early stage of the pandemic, HFNC was considered contraindicated for fear of aerosol spread, which was later proven not true in April (15). Thus, some patients put on ventilator before April probably could have been supported with HFNC. As such, the severity of hypoxemia in the no steroid group, and to a less extent in the methylprednisolone group, as measure by requirement of mechanical ventilation may have been an overestimation. However, the impact of this overestimation should be limited as our study period extended till end of July and most patients were treated after April. Besides, we have also used additional parameters, i.e., lowest PF ratio and highest PEEP, to further describe the severity of hypoxemia.

Our study is a single center retrospective study which needs verification in prospective, multi-center trials. However, our hospital is in one of the epicenters of the pandemic. As such, our patient cohort was sufficiently large and the patient population and the usual care including nursing were relatively homogenous. Although the data was collected retrospectively, the treatment regimens in our study for both steroids were specified from the beginning and consistently carried out throughout the study period. The amplitude of benefit due to dexamethasone matches closely to that reported in the RECOVERY Trial. The difference in survival benefit between the two steroids remained significant in patients on mechanical ventilation after 7 confounding factors were rigorously adjusted in the hazard ratio analysis. Thus, our results should add useful information to the optimal management of patients with severe COVID-19 pneumonia.

## CONCLUSIONS

The anti-inflammatory effects of corticosteroids have been associated with improved mortality outcomes in severe cases of COVID-19. The therapeutic benefits of corticosteroids are not limited to dexamethasone. Our results show that the higher anti-inflammatory potency of our methylprednisolone regimen can have greater mortality benefit in COVID-19 patients requiring mechanical ventilation.

## Data Availability

All the data are archived and available upone request

## ACKNOWLEDGEMENTS

We thank Drs. John Gruen and May Lee, as well as our many hospital colleagues for their assistance in data collection and patient care. We also thank the Internal Medicine residents (Drs. Dominic Engracia, Stefano Iantorno, and Ashil Panchal) for their valuable contribution in data collection for this study.

